# Community-based interventions to address depression in older adults: a systematic scoping review

**DOI:** 10.1101/2024.10.21.24315895

**Authors:** Laura Restrepo-Escudero, Maria Alejandra Jaimes, Isabela Arango, Sofia Santos, Valentina Ramírez, Daniel Uribe, Jenny Muñoz, Lina Maria González Ballesteros

**Affiliations:** Undergraduate Student, Faculty of Medicine, Pontificia Universidad Javeriana, Cra. 7 #40 - 62, Bogotá, Colombia; Doctor of Medicine, Faculty of Medicine, Pontificia Universidad Javeriana, Cra. 7 #40 - 62, Bogotá, Colombia; Psychiatry Resident, Department of Psychiatry and Mental Health, Faculty of Medicine, Pontificia Universidad Javeriana, Cra. 7 #40 - 62, Bogotá, Colombia; Master in Public Health, Saldarriaga Concha Foundation, Cl. 72 #7-64, Bogotá, Colombia; PhD (c) Clinical epidemiologist. Professor in the Department of Psychiatry and Mental Health, Faculty of Medicine, Pontificia Universidad Javeriana, Cra. 7 #40 - 62, Bogotá, Colombia; Saldarriaga Concha Foundation, Cl. 72 #7-64, Bogotá, Colombia

**Keywords:** Mental health, older people, public health strategies, public services

## Abstract

Depression is a prevalent psychiatric disorder among adults aged 65 and older, significantly impacting their well-being. With an aging global population, effective community-based interventions are vital to combat this issue. This review provides an overview of the characteristics of community-based interventions addressing depression in the elderly, identifying knowledge gaps by synthesizing current data. The search strategy entails a systematic database search. Results show that interventions have been effective in reducing depressive symptoms and enhancing social interaction, mainly those that include physical activity, social engagement, or mental health education. Success depends on factors such as participant engagement, adherence, and the sociocultural environment. Addressing these barriers requires a comprehensive understanding of local contexts and innovative service delivery approaches. Improving recruitment by overcoming cultural and logistical challenges could expand the reach and accessibility of these programs, thereby increasing their overall impact on older adults mental health and quality of life.

## Introduction

A significant social challenge of the twenty-first century is the aging of the world’s population. This will affect employment and financial markets, demand for housing, transportation, social security, family dynamics, and relationships between generations (1). The proportion of adults 65 years and beyond is rapidly increasing, signaling a projected rise from 10% in 2022 to 16% in 2050. Also, it is predicted to surpass the population of children under age 5 by 2050 and be nearly equal to children under age 12 (2).

Depression is a prevalent psychiatric disorder in this population, causing disability and reduced quality of life. It has been associated with several negative consequences, such as cognitive impairment, physical comorbidities, loneliness, premature mortality, and a poorer outcome of treatment for physical disorders (3–4). Depression prevalence among seniors has been examined, with estimates varying from 1% to 16% for major depression, 2% to 19% for minor depression, and 7.2% to 49% for depressive symptoms (5).

Various factors are associated with a higher prevalence of depression, like poor physical function, lack of a partner, and lower levels of education (5). It also seems to increase with age, regardless of the severity of the symptoms. Studies have reported a lower rate when excluding individuals living in institutions, as it appears to be higher in those hospitalized, institutionalized, or even in community settings (4). This highlights the importance of considering their living environment when investigating this disorder.

Community-based interventions are strategies applied within a defined local community to promote well-being among different populations, including the elderly. As they are highly heterogeneous and have different levels of efficiency, their impact changes depending on the characteristics of each community. Studies have shown that interventions focused on exercise, teaching new skills, encouraging community members to get involved in social activities, and using combination approaches can significantly lower depression scores in older adults (1).

Interventions incorporating physical activity, such as group fitness classes or individualized exercise programs, are associated with improved mood and reduced depression symptoms. Social isolation is a significant risk factor for depression in older adults, which can be mitigated through interventions promoting social engagement through group activities, volunteer work, or social support groups. Interventions combining multiple activities, such as social engagement, exercise, and cognitive stimulation, have been proven to be more effective in reducing depression than single-component interventions (6–7). Cognitive behavioral therapy (CBT) is another widely used and effective depression treatment. Community-based CBT interventions, delivered in any modality have been shown to effectively reduce depressive symptoms. They can be delivered through group sessions, not only addressing individual cognitive distortions and maladaptive behaviors but also promoting social interaction and peer support, critical components in mitigating the sense of isolation common among older adults (8).

Depression can also be addressed by improving psychosocial functioning or by promoting physical activity, which appears to have mood-related benefits for older adults (1). Approaches based on bibliotherapy increase self-management ability, and cognitive behavioral interventions delivered in this format of self-administered interventions have also been shown to improve mental health outcomes (9). It has been shown that implementing and reinforcing pleasant activities was one of the critical factors in making the interventions successful (9).

This review aims to provide an overview of the literature on community-based interventions for depression in this population, examining their features, goals, elements, activities, delivery, and dosage. We evaluated knowledge gaps in the literature and enhanced the understanding of this matter by analyzing the current status of research in these areas.

## Methodology

### Overview

This paper was conducted following the JBI methodology for scoping reviews (10) and designed as a systematic scoping review in line with the Preferred Items for Systematic Reviews and Meta-Analysis Guidelines extension for Scoping Reviews (PRISMA-ScR) (11). A systematic scoping review was conducted of the community-based interventions that have been developed for combating depression in adults 65 years and older. These include discussion groups, peer outreach programs, skill courses, and psychotherapeutic interventions, among others. In addition to describing the characteristics of these interventions (features, activities, mode, and frequency of delivery), mental health outcomes of interest include improvement of depressive symptoms or a decrease in the prevalence of depression in the elderly population in community settings. A review protocol was prepared for the guidance of the authors and was registered in the Open Science Framework: DOI: https://doi.org/10.17605/OSF.IO/MGCPZ.

### Eligibility criteria

All types of quantitative and qualitative studies were included if they reported community-based interventions that accomplished the improvement of depression symptoms and/or decreased the prevalence of depression in elderly adults. Information sources were restricted to community settings from any geographic region and cultural background. Studies were excluded if they evaluated services provided in healthcare settings, such as nursing homes or hospitals. Since this study aims to determine the effectiveness of interventions on depression as a primary psychiatric disorder, studies performed on patients with cognitive impairment, psychosis, and/or psychotic disorders were excluded. Opinion papers and unpublished studies were excluded. Finally, studies that lacked a quality review process and were not available in full text or a suitable format for analysis were excluded.

### Information sources

The information sources included peer-reviewed journals and preprint articles. We conducted an initial search in the PubMed database to find relevant articles on community-based interventions targeting depression in the elderly. We modified the keywords and index terms from the first search’s titles and abstracts to develop a customized search strategy for each database included. Considering the lack of literature on the topic, the search strategy was very sensible. Second, we conducted a systematic search of peer-reviewed and preprint literature in PubMed, EMBASE, Ovid, and PsycNet, without any language or time restrictions, from inception to July 2024, identifying sources reporting on mental health outcomes of interest, measured by improvement of depressive symptoms or decreased prevalence of depression. We reviewed articles that emerged from these searches and relevant references they cited. We included articles published in English and Spanish.

### Search strategy

The search of studies reporting community-based interventions for combating depression in the elderly was performed in PubMed, EMBASE, Ovid, and PsycNet using the following search strategy (shown as constructed in PubMed) without any time or language restrictions: (((“Aged”[Mesh]) OR (elderly OR “older adults” OR “older people”)) AND ((“Depression”[Mesh] OR “Depressive Disorder”[Mesh] OR “Depressive Disorder, Major”[Mesh]) OR (depres* OR “depressive disorder”)) AND ((“Community Mental Health Services”[Mesh]) OR (“community-based intervention” OR “community-based interventions” OR “community intervention” OR “community interventions”)) (See Appendix 1 for the complete search strategies).

### Selection of sources of evidence

All the included articles were uploaded into Rayyan QCRI software for duplicate removal. The titles and abstracts were screened by two reviewers (LRE, MAJ) to identify relevant studies. The full text of selected sources was assessed by seven reviewers (LRE, MAJ, IA, SS, OG, DU, VR) for eligibility as part of the systematic scoping review according to inclusion criteria. Disagreements between reviewers were resolved with an additional reviewer (LMG).

### Data charting process

Data collected from studies retrieved through the systematic search were extracted by the reviewers using piloted forms into a Microsoft Excel spreadsheet for analysis. Data charting was done independently and reviewed by one of the investigators (LRE).

### Data items

Data items extracted from all information sources regarding study characteristics included study design, context (e.g., target population, country, ethnicity, and setting), and sample characteristics (e.g., size, age, gender, and marital status). As to the community-based interventions, data items sought intervention characteristics in terms of components, activities, mode, and doses of delivery (referred to as frequency and duration). We classified and grouped the interventions based on the services they provided and then narratively summarized them. Mental health outcomes data items included tools for screening and diagnosing depression and/or depressive symptoms, along with any changes in the outcome following the implementation of the intervention. These changes were measured through differences in total questionnaire scores or by evaluating clinical progress through structured clinical interviews. We also collected qualitative data on factors associated with the implementation process, categorized them as ‘positively associated with implementation’ and ‘negatively associated with implementation’, and included them in the narrative summary. These determinants were thematically analyzed and grouped into various categories, such as ‘financial support’, ‘ability to tailor the intervention’,’staff training and supervision’, and ‘local staff’. Quantitative and qualitative data on the strengths and weaknesses of the interventions were collected and narratively summarized.

### Synthesis of results

We grouped the studies based on the type of intervention under evaluation. Information regarding the interventions’ characteristics was tabulated and summarized in the text. Categorized determinants associated with the implementation of different programs were identified for individual studies and outlined in tables and text. We narratively summarized the strengths and weaknesses of the studies. Quantitative meta-analysis was not performed due to high heterogeneity among studies.

## Results

### Selection of sources of evidence

The systematic search for community-based interventions addressing depression turned up 1109 records, of which 932 were screened. We included 57 studies for full-text screening after excluding 875 papers based on title and/or abstract. However, two articles (abstracts from scientific posters and unavailable journals) did not have full text, which left 55 final studies awaiting eligibility assessments. 13 studies were not conducted exclusively on elders aged above 65 years old or populations without dementia or psychosis; nine papers had an inadequate study focus, nine articles did not report measures of depression, and two were in languages unknown to authors (Chinese and Japanese), resulting in the inclusion of 23 studies (Fig 1).

**Figure 1:**
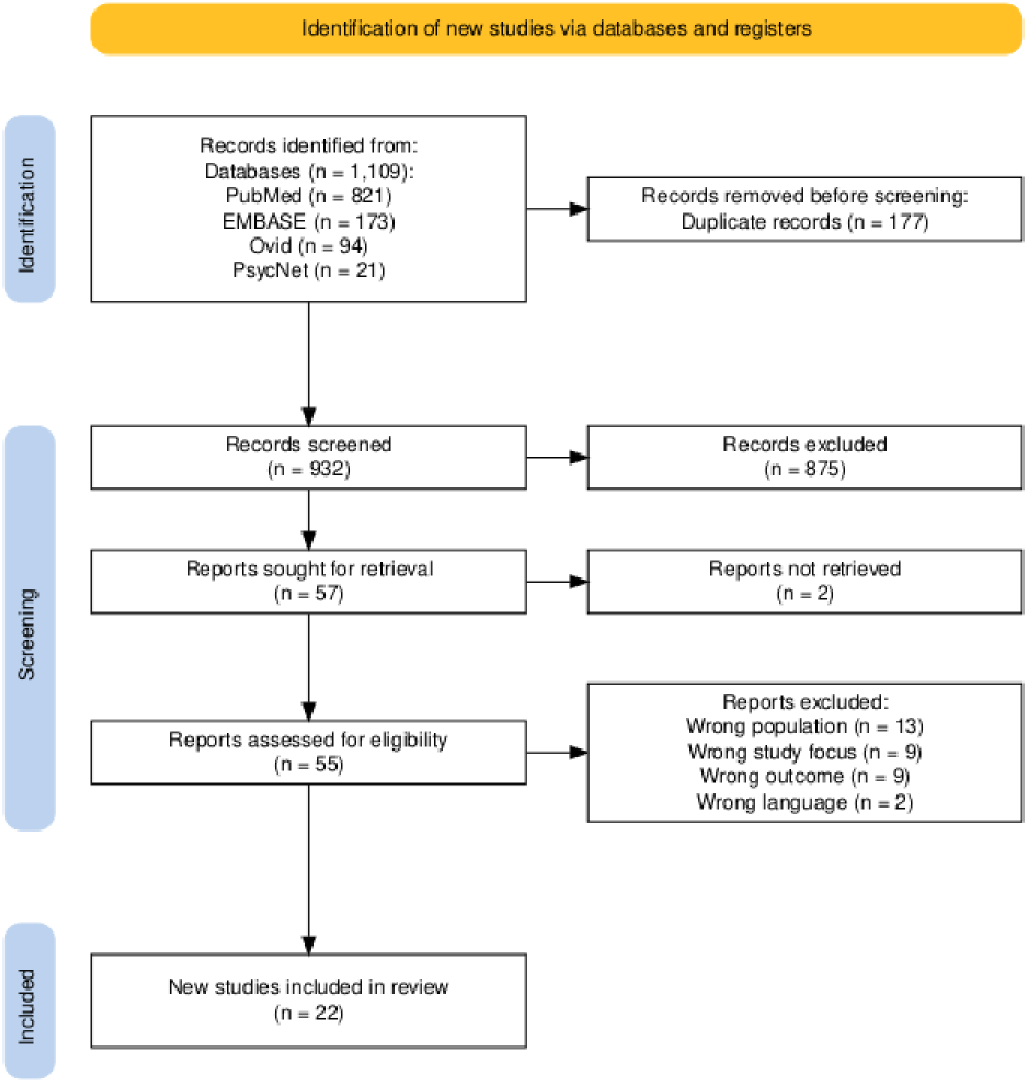
A flow diagram for selecting evidence sources.

### Characteristics of individual sources of evidence

Reviewed studies include a variety of approaches aimed at intervening in specific populations, primarily older adults, using methods in different cultural and geographical contexts. The characteristics of the 23 studies reviewed are presented in Table 1. Most studies were conducted in the United States, South Korea, and Japan. They employed various methodological designs, including mixed-methods longitudinal studies, randomized pilot trials, systematic reviews, and quasi-experimental cohort studies, with most studies (n=9) designed as randomized control trials (RCT). Researchers conducted studies in diverse cultural, social, and geographical contexts, including urban settings with low-income older adults (12) and rural areas in South Korea (13) and Japan (14). The samples vary in size, age, gender, and marital status. Populations included older adults from different ethnicities (African-American, Latin, Asian, Black, Indigenous, Caucasian, Hispanic), and the number of participants ranged from 6 to 1200. The studies were published between 1991 and 2023.

**Table 1:**
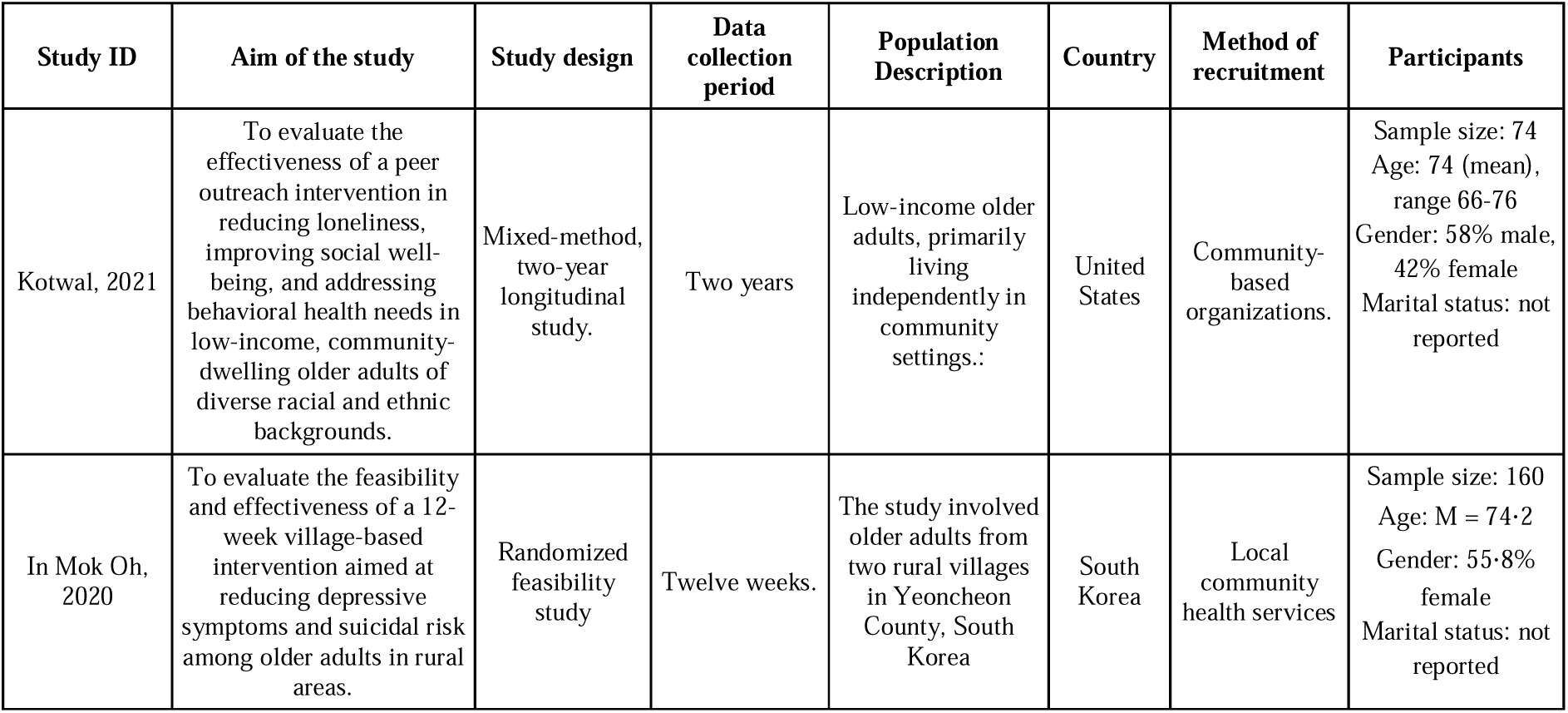
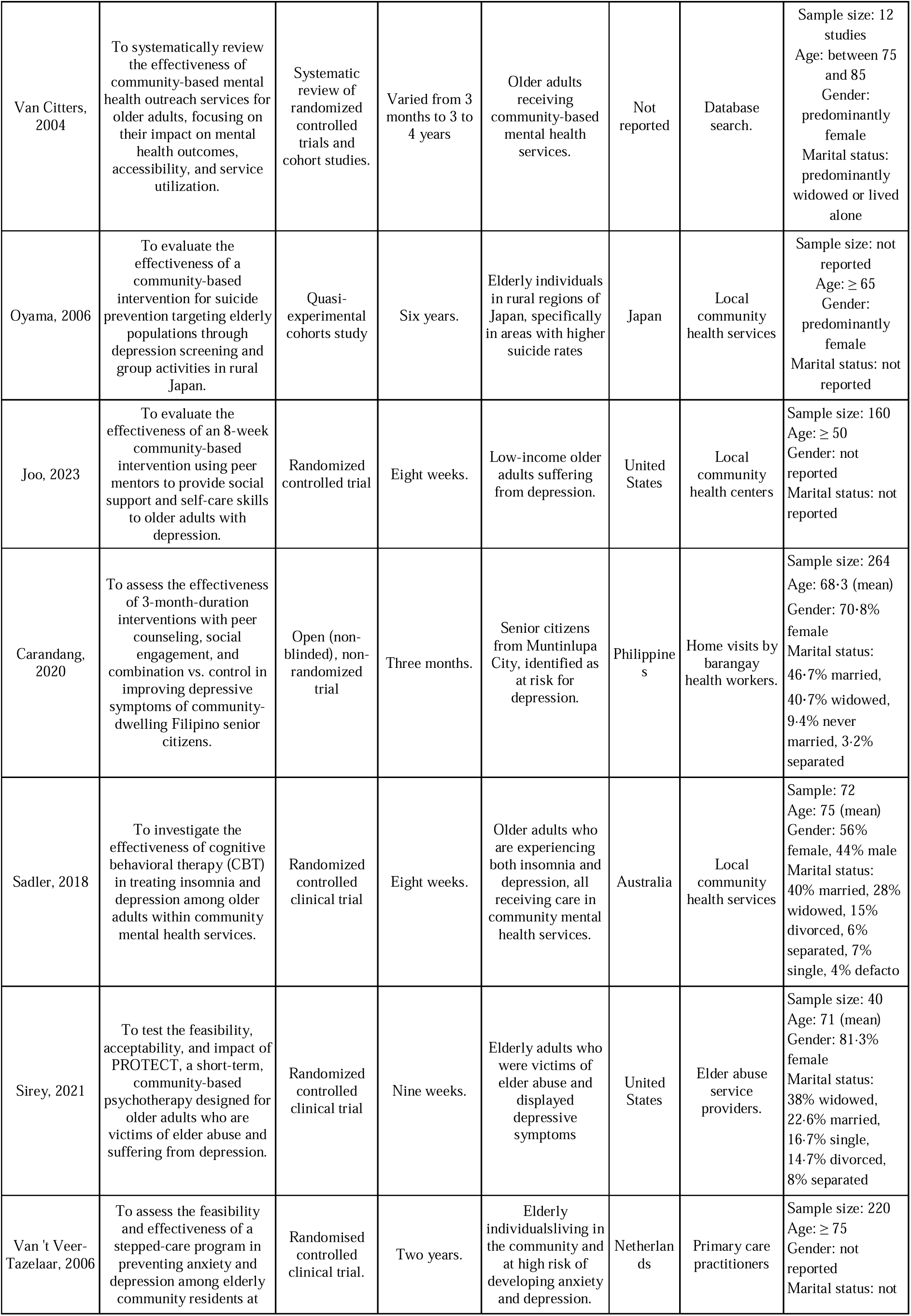
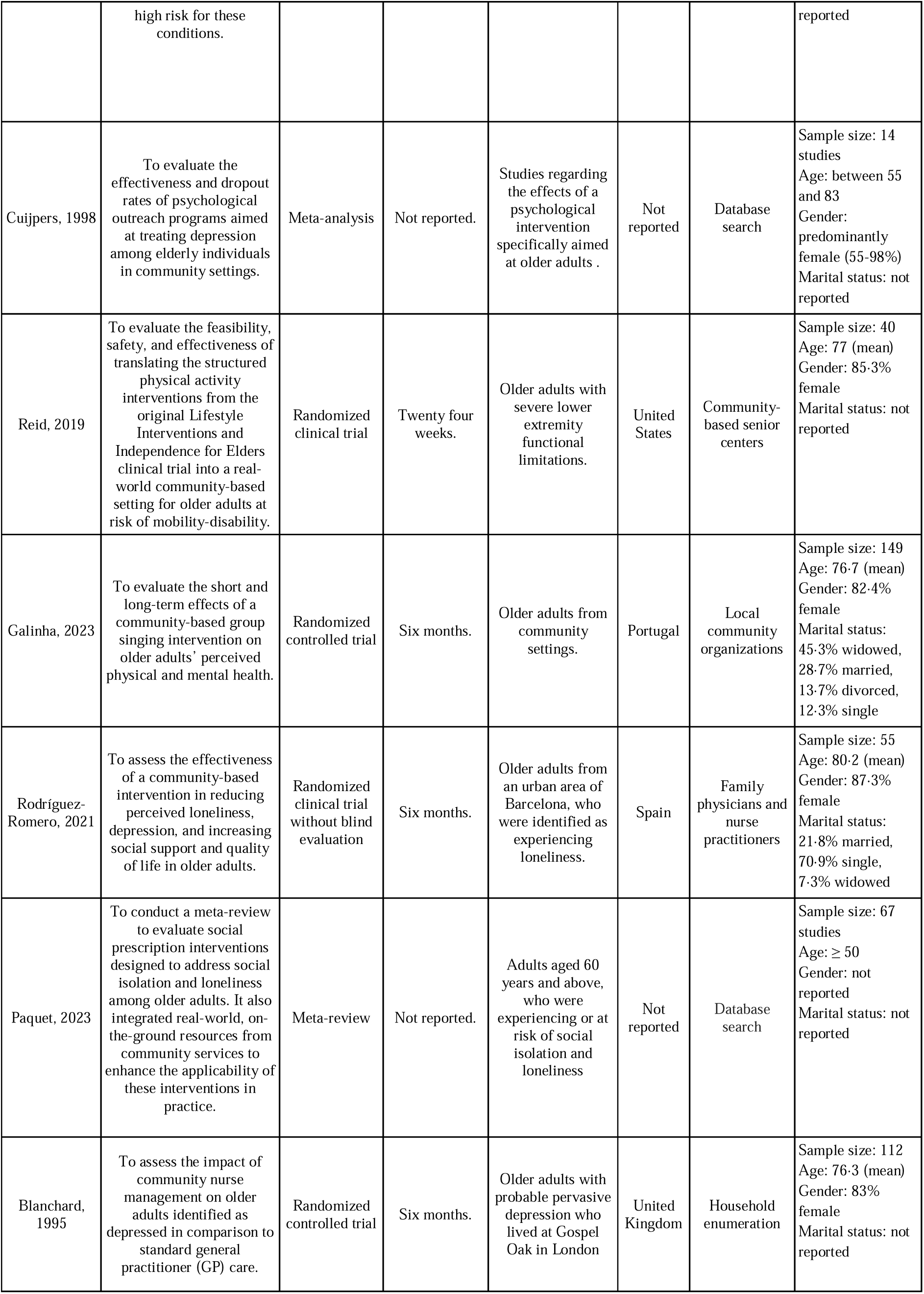
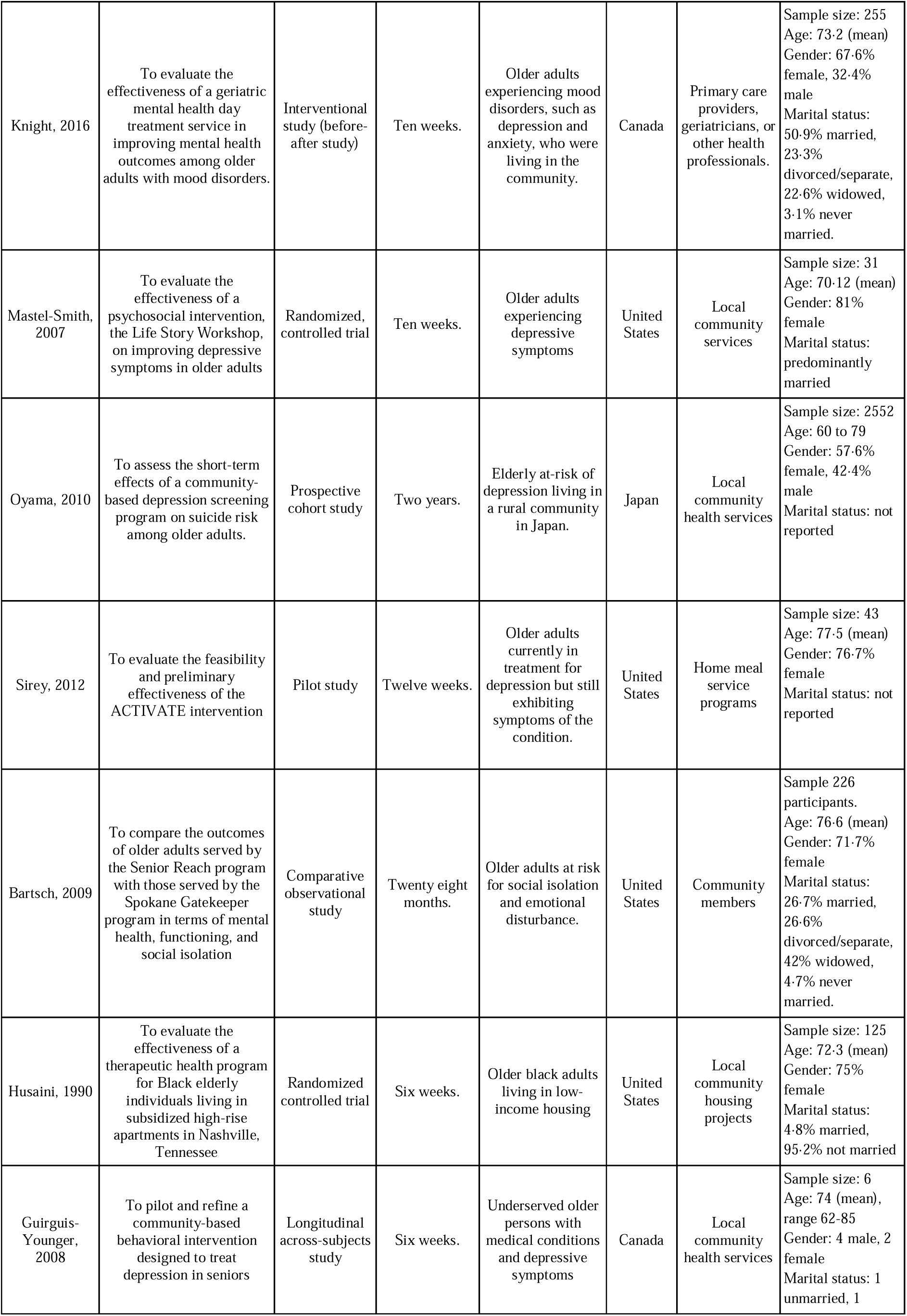
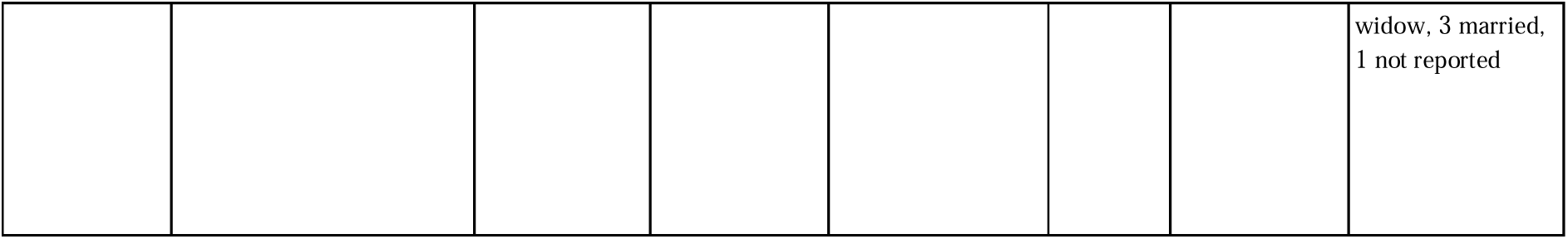
Characteristics of the studies reviewed.

### Synthesis of results

#### Types of interventions

Interventions were classified into personal contact, discussion groups, social engagement, skill course screening, and multicomponent interventions, which were programs that did not focus on a single type of intervention. The 23 studies included were grouped according to the intervention delivered, and their characteristics were described in Table 2. Personal contact interventions focus on social support through peer matching (12). Multicomponent interventions combine activities to address multiple factors, such as personal contact, social events, and exercise (13). Discussion-based and social engagement interventions promote social interaction and reduce loneliness (15). The activities performed include peer matching, home visits, and skills training (12, 16).

**Table 2:** Characteristics of the different types of interventions identified.

| Type of intervention | Components/Activities | Delivery mode | Doses Described |
| --- | --- | --- | --- |
| Discussion groups<br>(3 studies/reports)<br>Groups focused around discussing certain topics | 1. Reflection, writing, and sharing stories about their lived and current lives (Mastel-Smith, 2007)<br>2. Group discussions centered around seven modules (Husaini, 1990)<br>3. Group therapy (Sadler, 2018 ) | - Face-to-face sessions<br>- Groups of 10 people<br>- Lead by trained instructors | - 10 sessions once a week over an 11-week period<br>- Two weekly sessions of three hours each over a six week period<br>- One session (60 to 90 minutes long) per week over an eight week long period |
| Personal contact<br>( 2 studies/1 report)<br>Promotes contact with another person | 1. Therapy sessions in which an increase in social network could be requested if it was deemed necessary (Blanchard, 1995) | - Face-to-face sessions | - 10 visits, each of 45 minutes over a three-month period |
| Social engagement<br>(1 study / 1 report)<br>Engagement with a new group of people in an activity | 1. Group singing sessions (Galinha, 2023)<br>2. Group based social activities (Paquet, 2023) | - Face-to-face sessions | - 34 sessions of two hours over a four month period |
| Skill course<br>(1 study)<br>Courses delivered intended to help develop participants' personal skills. | 1. Lectures centered around certain topics like depression and anxiety, reading materials were provided on problem solving strategies, motivational techniques, and depression care options and their efficacy. After initial training, role-play and experience are encouraged | - Face-to-face sessions<br>- Lead by trained instructors (social workers) | - At least two, 30-minute meetings |
| Screening<br>(1 study)<br>Interventions that focused on implementing depression screening tools. | 1. An anonymous survey by random sample in the entire intervention region was delivered and a depression screening with follow-up by a psychiatrist | - Self administered | - Two-year implementation period |
| Discussion groups and personal contact<br>(4 studies/reports)<br>Interventions that implemented both personal contact and discussion groups | 1. Local visits done by fellow residents, who identified worsening symptoms and referred them to medical professionals if needed. They also conducted group sessions held at the community hall for enhancing interpersonal networks (In Mok Oh, 2020)<br>2. Problem solving therapy (Van Citters, 2004)<br>3. Workshops for psychoeducation held together with group activity programs in the intervention area (Oyama, 2006)<br>4. Peers provided social support and self-care skills to depressed older adults (Joo, 2023) | - Groups of 10 to 12 people<br>- One to one contact<br>- Staff involved social workers, nurses, care managers, physicians and residential staff | - Programs lasted from 12 weeks to four years |
| Skill course and personal contact<br>(4 studies/reports)<br>Interventions that implemented both skill courses and personal contact | 1. The intervention group received a stepped care program. which included psychoeducation, self-help materials, and personalized support (Van 't Veer-Tazelaar, 2006)<br>2. Community outings focused on behavioral activation and social skills, therapy sessions focused around reinforcing the group - based interventions, cognitive-behavioral therapy, and interpersonal psychotherapy modalities, exercise, psychoeducation and skill-development groups, and individual therapy sessions (Knight, 2016)<br>3. Self-administered bibliotherapy intervention based on a cognitive-behavioral approach. Written material with examples and exercises were developed (Guirguis-Younger, 2008)<br>4. Activities including educational workshops, mind- fulness, yoga, walking and visits to urban gardens (Rodríguez-Romero, 2021) | - Self-administered<br>- One to one contact<br>- Staff involved trained nurses | - Programs lasted from six weeks to two years |
| Multicomponent<br>(3 studies/reports)<br>Programs that did not<br>focus on a single type of<br>intervention | 1. Peer Counseling sessions conducted by senior volunteers, social events that included dancing, educational talks, group discussions, interactive games, and karaoke (Carandang, 2020)<br>2. Aerobic walking, strength training, flexibility exercises, balance training sessions , alongside workshops on topics relevant to older adults (Reid, 2019)<br>3. Community members were trained to identify and refer at risk older adults (Bartsch, 2009) | - Face-to-face sessions<br>- Interventions were facilitated by staff which included volunteers from the community and health providers<br>- Call centers | - Sessions once or twice a week<br>- Programs lasted from three months up to two years |

Trained instructors usually developed interventions; however, sometimes, studies involved the community for the implementation of the programs, with peer outreach approaches, local community volunteers, and partnerships with local organizations. Active community involvement helps increase elderly participation. Educational and social activity programs tailored for elderly women showed remarkable success in reducing suicide rates by 74% (14). Such community-driven efforts created a sense of belonging and trust, which encourages sustained participation and contributes to overall success (12). Other studies collaborated with trusted community organizations and interventions were delivered by their staff, which included social workers, nurses, and care managers (9, 17, 18). These organizations helped recruit participants, disseminate mental health resources in a non-stigmatizing and familiar manner, and facilitate the customization and delivery of interventions to address specific needs and provide a more accessible and feasible form of therapy (9, 19–20).

The effectiveness of interventions was evaluated using standardized scales to measure the depression levels among participants. Commonly used tools include the geriatric depression scale (GDS) and the Patient Health Questionnaire (PHQ-9), both of which are well-regarded for their reliability. In older adults, the majority of interventions reported significant improvements in depression scores that indicate a positive impact on the participants’ perceptions of wellness and mental health. For instance, programs that incorporated personal contact and social engagement, like peer matching and home visits, consistently demonstrated a reduction in depressive symptoms (9, 17–18, 20–24).

A few interventions did not meet the expected outcomes, highlighting the complexity of addressing mental health in this population. In Mok Oh et al., although their focus was reducing depressive symptoms in rural communities, depression scores did not improve as expected (13). Similarly, in the multi-component approach shown by Oyama et al., where they aimed to prevent suicide in elderly populations, even though it was effective regarding engagement and participation, it did not have substantial reductions in depression (14). Factors like deferring levels of participant engagement, specific needs of the target demographic, or duration of the intervention may account for these variations in effectiveness (9). Nevertheless, the overall evidence strongly supports the effectiveness of community-based approaches in improving older adults’ perceptions of mental health and well-being.

#### The factors that determine how an intervention is implemented

Many socio-cultural, financial, and geographical contexts influence the implementation of health interventions, particularly those targeting mental health. Table 3 presents the determinants that facilitate or hinder the success of interventions found in the 23 studies, and Table 4 summarizes them. Most studies did not systematically identify these factors, favoring the possibility of publication bias and missing data in the analysis.

**Table 3:**
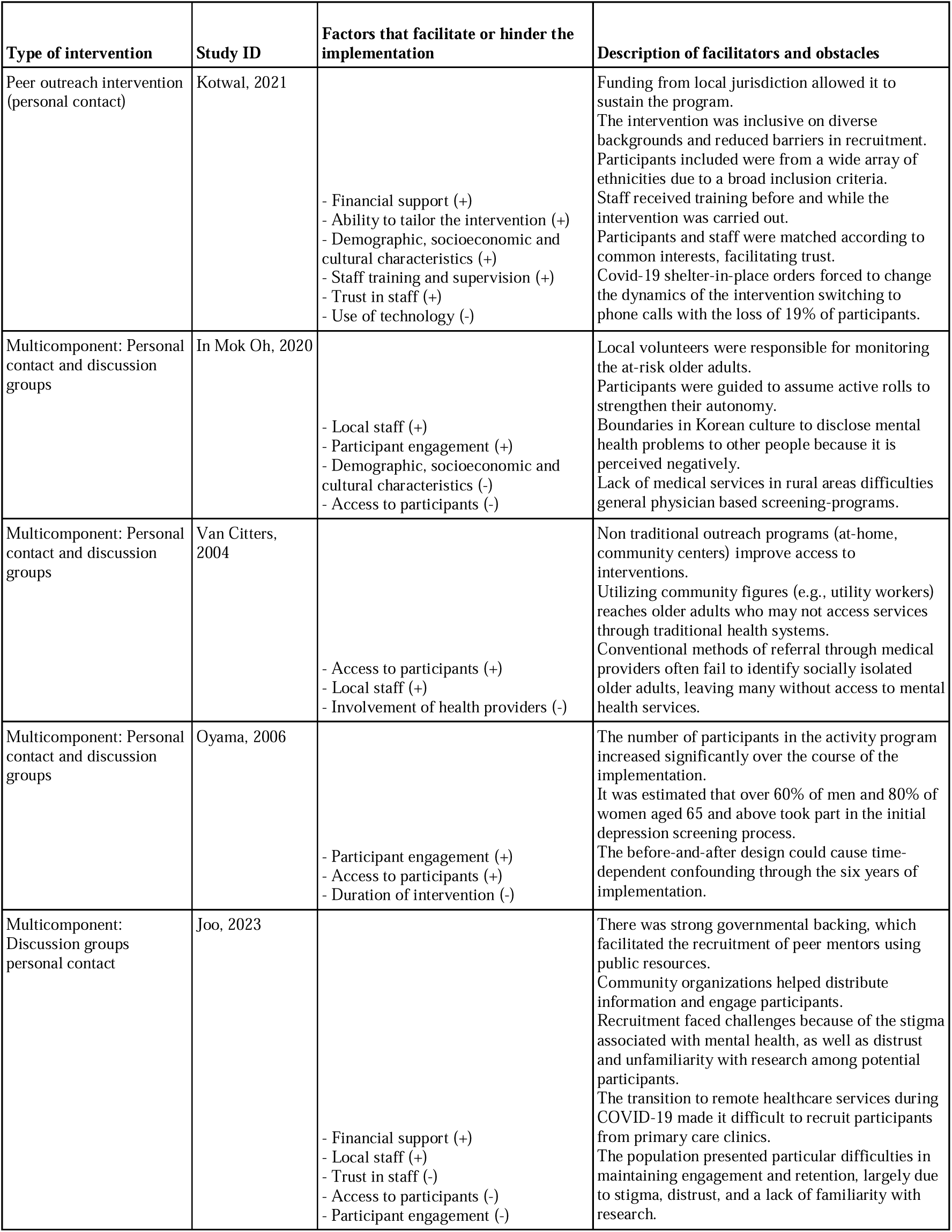
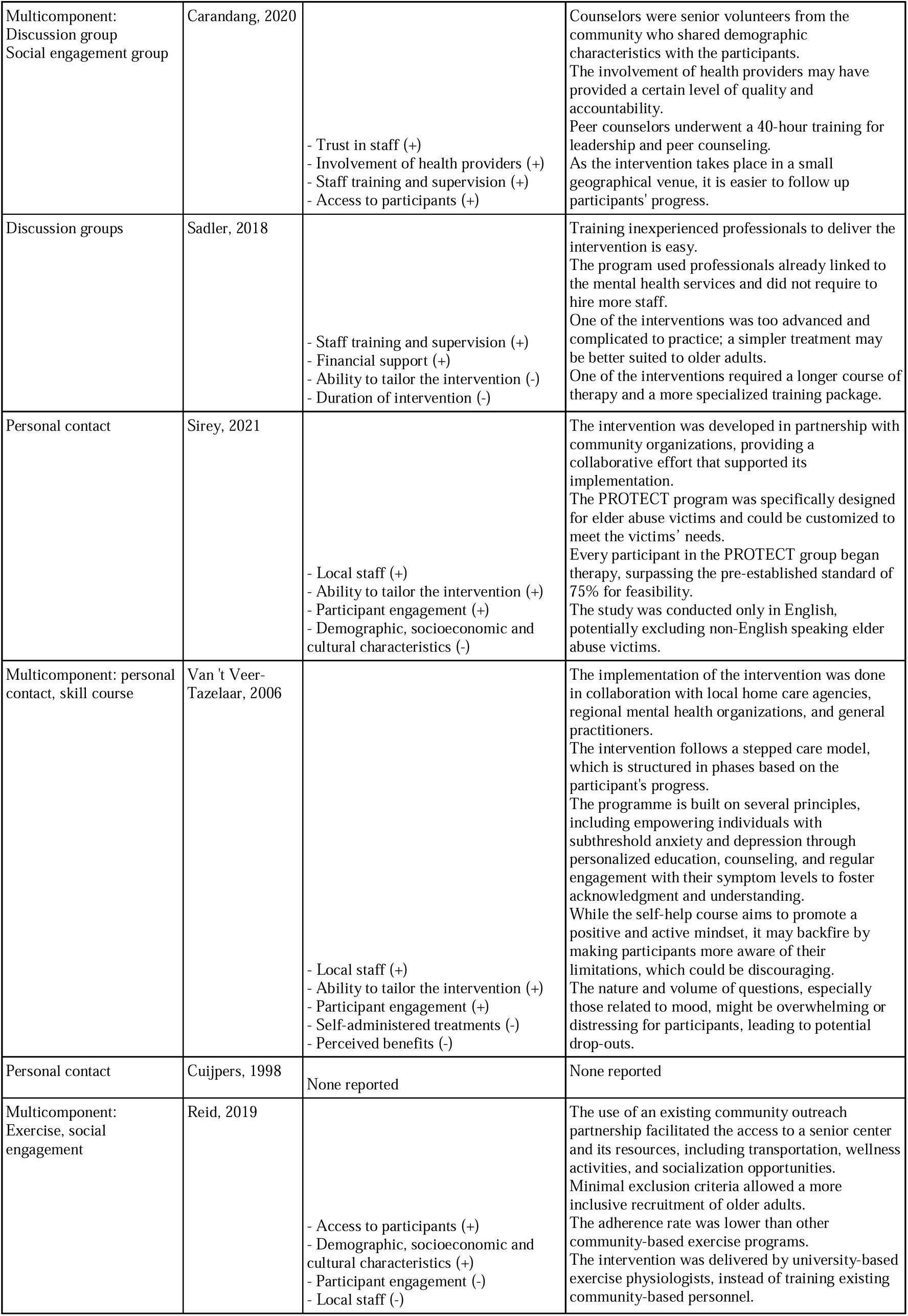
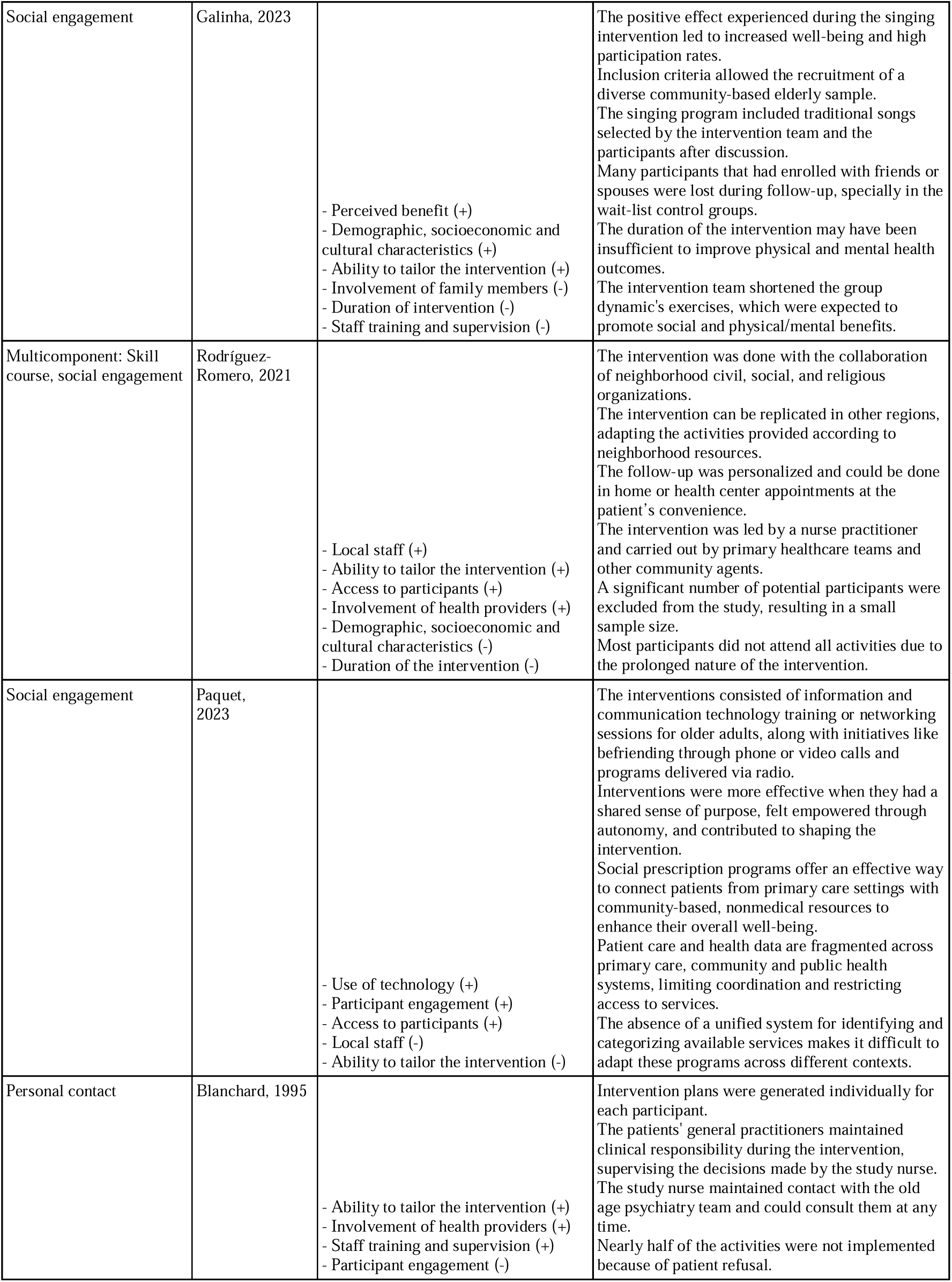
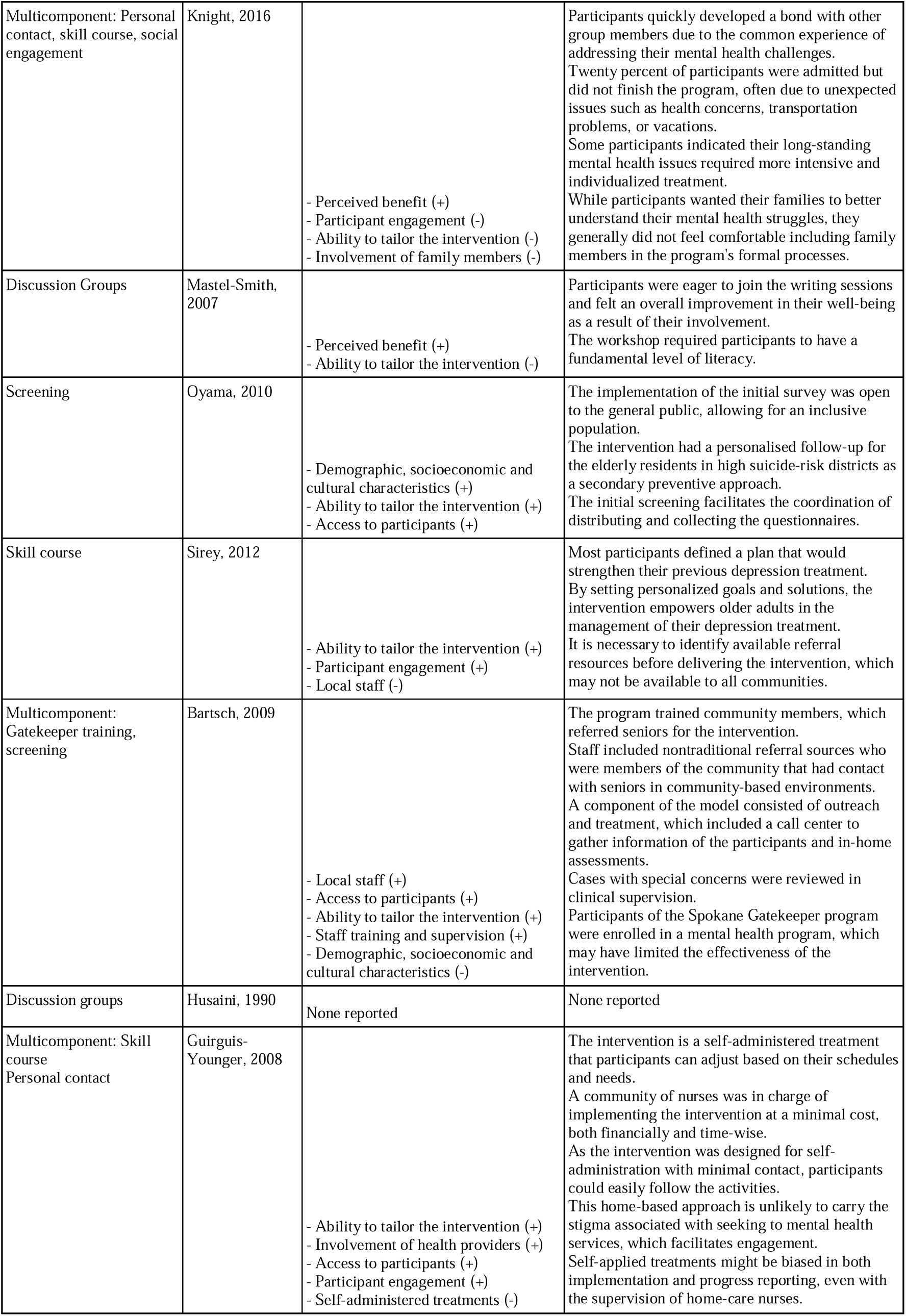
Factors that facilitate or hinder the implementation of the interventions.

| Type of intervention | Study ID | Factors that facilitate or hinder the implementation | Description of facilitators and obstacles |
| --- | --- | --- | --- |
| Peer outreach intervention (personal contact) | Kotwal, 2021 | - Financial support (+)<br>- Ability to tailor the intervention (+)<br>- Demographic, socioeconomic and cultural characteristics (+)<br>- Staff training and supervision (+)<br>- Trust in staff (+)<br>- Use of technology (-) | Funding from local jurisdiction allowed it to sustain the program.<br>The intervention was inclusive on diverse backgrounds and reduced barriers in recruitment. Participants included were from a wide array of ethnicities due to a broad inclusion criteria. Staff received training before and while the intervention was carried out.<br>Participants and staff were matched according to common interests, facilitating trust.<br>Covid-19 shelter-in-place orders forced to change the dynamics of the intervention switching to phone calls with the loss of 19% of participants. |
| Multicomponent: Personal contact and discussion groups | In Mok Oh, 2020 | - Local staff (+)<br>- Participant engagement (+)<br>- Demographic, socioeconomic and cultural characteristics (-)<br>- Access to participants (-) | Local volunteers were responsible for monitoring the at-risk older adults.<br>Participants were guided to assume active roles to strengthen their autonomy.<br>Boundaries in Korean culture to disclose mental health problems to other people because it is perceived negatively.<br>Lack of medical services in rural areas difficulties general physician based screening-programs. |
| Multicomponent: Personal contact and discussion groups | Van Citters, 2004 | - Access to participants (+)<br>- Local staff (+)<br>- Involvement of health providers (-) | Non traditional outreach programs (at-home, community centers) improve access to interventions.<br>Utilizing community figures (e.g., utility workers) reaches older adults who may not access services through traditional health systems.<br>Conventional methods of referral through medical providers often fail to identify socially isolated older adults, leaving many without access to mental health services. |
| Multicomponent: Personal contact and discussion groups | Oyama, 2006 | - Participant engagement (+)<br>- Access to participants (+)<br>- Duration of intervention (-) | The number of participants in the activity program increased significantly over the course of the implementation.<br>It was estimated that over 60% of men and 80% of women aged 65 and above took part in the initial depression screening process.<br>The before-and-after design could cause time-dependent confounding through the six years of implementation. |
| Multicomponent: Discussion groups personal contact | Joo, 2023 | - Financial support (+)<br>- Local staff (+)<br>- Trust in staff (-)<br>- Access to participants (-)<br>- Participant engagement (-) | There was strong governmental backing, which facilitated the recruitment of peer mentors using public resources.<br>Community organizations helped distribute information and engage participants.<br>Recruitment faced challenges because of the stigma associated with mental health, as well as distrust and unfamiliarity with research among potential participants.<br>The transition to remote healthcare services during COVID-19 made it difficult to recruit participants from primary care clinics.<br>The population presented particular difficulties in maintaining engagement and retention, largely due to stigma, distrust, and a lack of familiarity with research. |
| Multicomponent:<br>Discussion group<br>Social engagement group | Carandang, 2020 | - Trust in staff (+)<br>- Involvement of health providers (+)<br>- Staff training and supervision (+)<br>- Access to participants (+) | Counselors were senior volunteers from the community who shared demographic characteristics with the participants.<br>The involvement of health providers may have provided a certain level of quality and accountability.<br>Peer counselors underwent a 40-hour training for leadership and peer counseling.<br>As the intervention takes place in a small geographical venue, it is easier to follow up participants' progress. |
| Discussion groups | Sadler, 2018 | - Staff training and supervision (+)<br>- Financial support (+)<br>- Ability to tailor the intervention (-)<br>- Duration of intervention (-) | Training inexperienced professionals to deliver the intervention is easy.<br>The program used professionals already linked to the mental health services and did not require to hire more staff.<br>One of the interventions was too advanced and complicated to practice; a simpler treatment may be better suited to older adults.<br>One of the interventions required a longer course of therapy and a more specialized training package. |
| Personal contact | Sirey, 2021 | - Local staff (+)<br>- Ability to tailor the intervention (+)<br>- Participant engagement (+)<br>- Demographic, socioeconomic and cultural characteristics (-) | The intervention was developed in partnership with community organizations, providing a collaborative effort that supported its implementation.<br>The PROTECT program was specifically designed for elder abuse victims and could be customized to meet the victims' needs.<br>Every participant in the PROTECT group began therapy, surpassing the pre-established standard of 75% for feasibility.<br>The study was conducted only in English, potentially excluding non-English speaking elder abuse victims. |
| Multicomponent: personal contact, skill course | Van 't Veer-Tazelaar, 2006 | - Local staff (+)<br>- Ability to tailor the intervention (+)<br>- Participant engagement (+)<br>- Self-administered treatments (-)<br>- Perceived benefits (-) | The implementation of the intervention was done in collaboration with local home care agencies, regional mental health organizations, and general practitioners.<br>The intervention follows a stepped care model, which is structured in phases based on the participant's progress.<br>The programme is built on several principles, including empowering individuals with subthreshold anxiety and depression through personalized education, counseling, and regular engagement with their symptom levels to foster acknowledgment and understanding.<br>While the self-help course aims to promote a positive and active mindset, it may backfire by making participants more aware of their limitations, which could be discouraging.<br>The nature and volume of questions, especially those related to mood, might be overwhelming or distressing for participants, leading to potential drop-outs. |
| Personal contact | Cuijpers, 1998 | None reported | None reported |
| Multicomponent:<br>Exercise, social engagement | Reid, 2019 | - Access to participants (+)<br>- Demographic, socioeconomic and cultural characteristics (+)<br>- Participant engagement (-)<br>- Local staff (-) | The use of an existing community outreach partnership facilitated the access to a senior center and its resources, including transportation, wellness activities, and socialization opportunities.<br>Minimal exclusion criteria allowed a more inclusive recruitment of older adults.<br>The adherence rate was lower than other community-based exercise programs.<br>The intervention was delivered by university-based exercise physiologists, instead of training existing community-based personnel. |
| Social engagement | Galinha, 2023 | - Perceived benefit (+)<br>- Demographic, socioeconomic and cultural characteristics (+)<br>- Ability to tailor the intervention (+)<br>- Involvement of family members (-)<br>- Duration of intervention (-)<br>- Staff training and supervision (-) | The positive effect experienced during the singing intervention led to increased well-being and high participation rates.<br>Inclusion criteria allowed the recruitment of a diverse community-based elderly sample.<br>The singing program included traditional songs selected by the intervention team and the participants after discussion.<br>Many participants that had enrolled with friends or spouses were lost during follow-up, specially in the wait-list control groups.<br>The duration of the intervention may have been insufficient to improve physical and mental health outcomes.<br>The intervention team shortened the group dynamic's exercises, which were expected to promote social and physical/mental benefits. |
| Multicomponent: Skill course, social engagement | Rodríguez-Romero, 2021 | - Local staff (+)<br>- Ability to tailor the intervention (+)<br>- Access to participants (+)<br>- Involvement of health providers (+)<br>- Demographic, socioeconomic and cultural characteristics (-)<br>- Duration of the intervention (-) | The intervention was done with the collaboration of neighborhood civil, social, and religious organizations.<br>The intervention can be replicated in other regions, adapting the activities provided according to neighborhood resources.<br>The follow-up was personalized and could be done in home or health center appointments at the patient's convenience.<br>The intervention was led by a nurse practitioner and carried out by primary healthcare teams and other community agents.<br>A significant number of potential participants were excluded from the study, resulting in a small sample size.<br>Most participants did not attend all activities due to the prolonged nature of the intervention. |
| Social engagement | Paquet, 2023 | - Use of technology (+)<br>- Participant engagement (+)<br>- Access to participants (+)<br>- Local staff (-)<br>- Ability to tailor the intervention (-) | The interventions consisted of information and communication technology training or networking sessions for older adults, along with initiatives like befriending through phone or video calls and programs delivered via radio.<br>Interventions were more effective when they had a shared sense of purpose, felt empowered through autonomy, and contributed to shaping the intervention.<br>Social prescription programs offer an effective way to connect patients from primary care settings with community-based, nonmedical resources to enhance their overall well-being.<br>Patient care and health data are fragmented across primary care, community and public health systems, limiting coordination and restricting access to services.<br>The absence of a unified system for identifying and categorizing available services makes it difficult to adapt these programs across different contexts. |
| Personal contact | Blanchard, 1995 | - Ability to tailor the intervention (+)<br>- Involvement of health providers (+)<br>- Staff training and supervision (+)<br>- Participant engagement (-) | Intervention plans were generated individually for each participant.<br>The patients' general practitioners maintained clinical responsibility during the intervention, supervising the decisions made by the study nurse.<br>The study nurse maintained contact with the old age psychiatry team and could consult them at any time.<br>Nearly half of the activities were not implemented because of patient refusal. |
| Multicomponent: Personal contact, skill course, social engagement | Knight, 2016 | - Perceived benefit (+)<br>- Participant engagement (-)<br>- Ability to tailor the intervention (-)<br>- Involvement of family members (-) | Participants quickly developed a bond with other group members due to the common experience of addressing their mental health challenges. Twenty percent of participants were admitted but did not finish the program, often due to unexpected issues such as health concerns, transportation problems, or vacations.<br>Some participants indicated their long-standing mental health issues required more intensive and individualized treatment.<br>While participants wanted their families to better understand their mental health struggles, they generally did not feel comfortable including family members in the program's formal processes. |
| Discussion Groups | Mastel-Smith, 2007 | - Perceived benefit (+)<br>- Ability to tailor the intervention (-) | Participants were eager to join the writing sessions and felt an overall improvement in their well-being as a result of their involvement.<br>The workshop required participants to have a fundamental level of literacy. |
| Screening | Oyama, 2010 | - Demographic, socioeconomic and cultural characteristics (+)<br>- Ability to tailor the intervention (+)<br>- Access to participants (+) | The implementation of the initial survey was open to the general public, allowing for an inclusive population.<br>The intervention had a personalised follow-up for the elderly residents in high suicide-risk districts as a secondary preventive approach.<br>The initial screening facilitates the coordination of distributing and collecting the questionnaires. |
| Skill course | Sirey, 2012 | - Ability to tailor the intervention (+)<br>- Participant engagement (+)<br>- Local staff (-) | Most participants defined a plan that would strengthen their previous depression treatment. By setting personalized goals and solutions, the intervention empowers older adults in the management of their depression treatment.<br>It is necessary to identify available referral resources before delivering the intervention, which may not be available to all communities. |
| Multicomponent: Gatekeeper training, screening | Bartsch, 2009 | - Local staff (+)<br>- Access to participants (+)<br>- Ability to tailor the intervention (+)<br>- Staff training and supervision (+)<br>- Demographic, socioeconomic and cultural characteristics (-) | The program trained community members, which referred seniors for the intervention.<br>Staff included nontraditional referral sources who were members of the community that had contact with seniors in community-based environments.<br>A component of the model consisted of outreach and treatment, which included a call center to gather information of the participants and in-home assessments.<br>Cases with special concerns were reviewed in clinical supervision.<br>Participants of the Spokane Gatekeeper program were enrolled in a mental health program, which may have limited the effectiveness of the intervention. |
| Discussion groups | Husaini, 1990 | None reported | None reported |
| Multicomponent: Skill course<br>Personal contact | Guirguis-Younger, 2008 | - Ability to tailor the intervention (+)<br>- Involvement of health providers (+)<br>- Access to participants (+)<br>- Participant engagement (+)<br>- Self-administered treatments (-) | The intervention is a self-administered treatment that participants can adjust based on their schedules and needs.<br>A community of nurses was in charge of implementing the intervention at a minimal cost, both financially and time-wise.<br>As the intervention was designed for self-administration with minimal contact, participants could easily follow the activities.<br>This home-based approach is unlikely to carry the stigma associated with seeking to mental health services, which facilitates engagement.<br>Self-applied treatments might be biased in both implementation and progress reporting, even with the supervision of home-care nurses. |

**Table 4:**
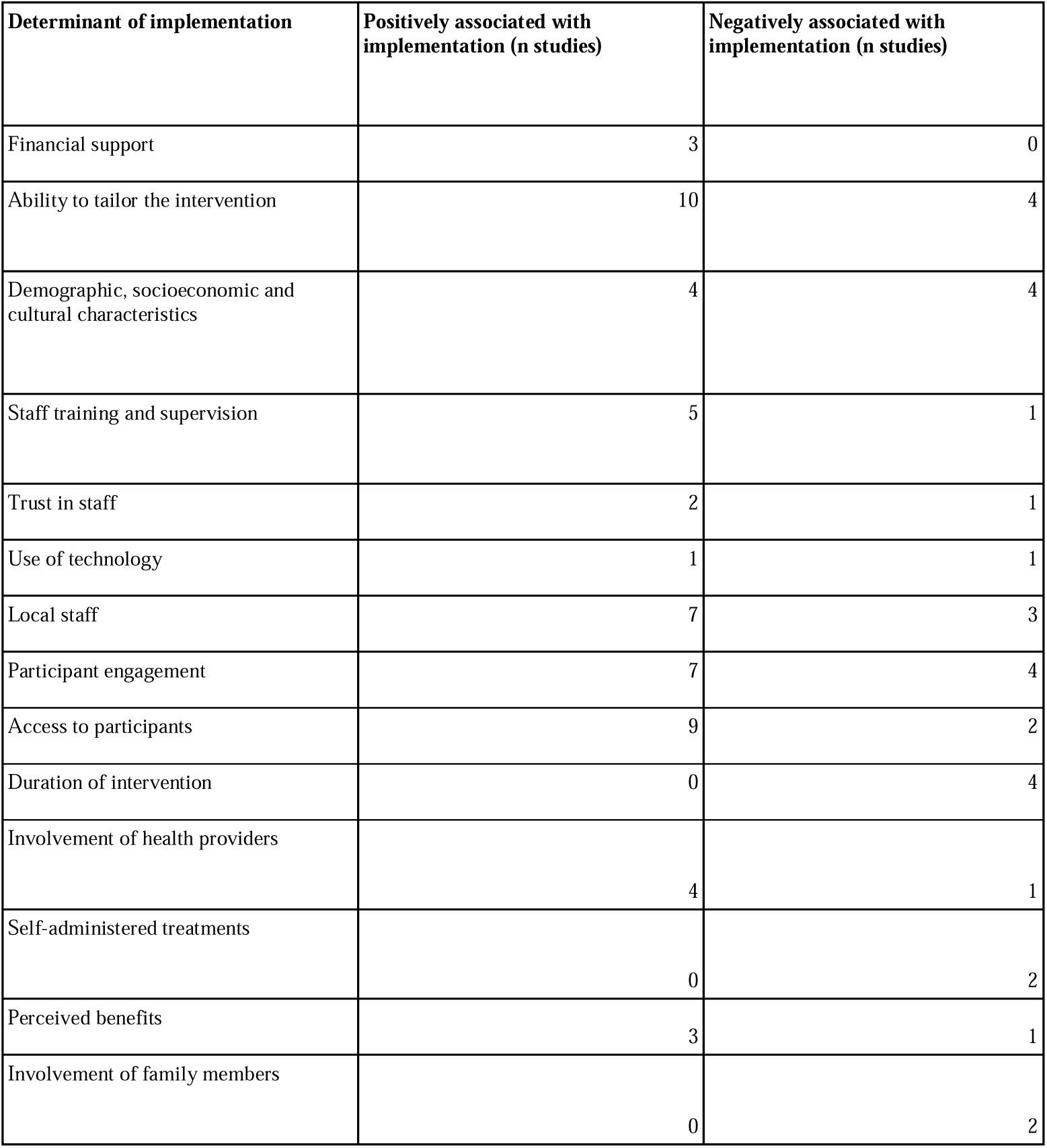
Determinants and how they were associated with the implementation of the intervention.

| Determinant of implementation | Positively associated with implementation (n studies) | Negatively associated with implementation (n studies) |
| --- | --- | --- |
| Financial support | 3 | 0 |
| Ability to tailor the intervention | 10 | 4 |
| Demographic, socioeconomic and cultural characteristics | 4 | 4 |
| Staff training and supervision | 5 | 1 |
| Trust in staff | 2 | 1 |
| Use of technology | 1 | 1 |
| Local staff | 7 | 3 |
| Participant engagement | 7 | 4 |
| Access to participants | 9 | 2 |
| Duration of intervention | 0 | 4 |
| Involvement of health providers | 4 | 1 |
| Self-administered treatments | 0 | 2 |
| Perceived benefits | 3 | 1 |
| Involvement of family members | 0 | 2 |

One of the most recurrent factors facilitating the implementation of mental health interventions is the presence of community engagement with the involvement of local staff (13, 17–20). Partnering with community groups helps the programs provide a more accessible and feasible form of therapy (20). The ability to tailor interventions to participants’ needs and backgrounds is another important facilitator for increasing participant engagement, accessibility, and inclusion. These factors are independently associated with improved implementation strategies (9, 17, 25).

Financial support emerged as another factor facilitating the implementation of the intervention programs (12, 14). It facilitates recruitment, enables interventions to reach populations, and provides necessary resources to train the staff, simplifying implementation (26–27). Technology also provided tools that facilitated interventions and boosted engagement with older adults (28). Broad inclusion criteria allowed the recruitment of participants from underrepresented groups, increasing the feasibility of the interventions (29). The involvement of health providers alongside local staff contributed to building trust and accountability, maintaining clinical responsibility, and overall supervision, which facilitated the implementation of the interventions and greater participant engagement and follow-up (9, 23, 30).

Despite the evident benefits, several barriers still hinder the successful implementation of community-based interventions, which include cultural stigmas surrounding seeking mental health services. Many cultures perceive discussing mental health as shameful, thereby limiting their participation and efficacy (13). Interventions’ duration was another common setback, due to insufficient time to accomplish long-term objectives or because of the prolonged nature of decreased adherence (18, 25). The involvement of family members appeared as a barrier, mainly due to fear of disclosing mental health issues and fear of being allocated to a different intervention group (21, 25). Self-administered treatments also increased individuals’ awareness of their limitations, potentially leading to bias in the implementation and reporting of progress (9, 17).

Notwithstanding the aforesaid, other contextual factors influenced the adoption of many implementations. Sociocultural backgrounds include cultural barriers that limit participation, such as reluctance in South Korea to discuss mental health issues and social norms in Japan affecting the willingness to seek help (13–14). Such stigma in mental health poses a challenge to participants’ recruitment and acceptance of mental health care (12, 23, 26). Additionally, some studies were only carried out in English, excluding other ethnicities and backgrounds or lacking diversity with more women and white ethnicity representation (16, 20).

The geographical environment influences the implementation of interventions, with challenges in rural areas and a lack of medical services that hinder them in isolated populations (13). Rural areas present logistical difficulties, demanding innovative outreach methods to overcome geographical barriers (15). In rural areas, bringing services to people’s homes or community centers has been shown to reduce some of them (15). Rural areas, on the other hand, facilitate socialization among neighbors because of reduced physical distance (13).

Conversely, urban settings benefit from better infrastructure, which improves accessibility to participants, favors adherence to interventions, and promotes social contact with local community members (19, 21). However, the lack of comprehensive cost-effectiveness studies also limits the potential for scaling these interventions to larger populations (15).

#### The intervention’s strengths and weaknesses

One key strength across the studies was the emphasis on enhancing socialization. Through recreational activities, self-organized social groups, or health education sessions, many interventions reduced loneliness, fostering emotional well-being by building social connections (12–13, 21, 24). Other studies adopted a holistic approach, focusing on treating comorbidities, which improved depression symptoms (27).

Another strength is the flexibility of these interventions, which allowed them to meet the needs of each community. The multidisciplinary approach endorsed by many programs and the involvement of participants in the design and execution of activities enhanced the sustainability and relevance of these interventions (15, 18, 28), resulting in high participation and retention rates (14, 19, 27). A further strength lies in their cost-effectiveness, scalability, and safety. Programs depend on low-cost, self-administered activities, methods with little supervision, or the help of local volunteers. This implies that they are applicable in locations with limited resources (9, 24). The lack of safety concerns and the scalability of such interventions ensure their implementation across various contexts, including rural and urban environments, without the need for substantial financial investment and rigorous medical surveillance (27, 29).

Methodologically, many studies used comprehensive assessment tools to evaluate mental health outcomes. Researchers employed validated scales such as the GDS and PHQ-9 to monitor participants’ progress and other methods like structured clinical interviews for diagnosis, which improves the reliability of findings (16, 20).

Furthermore, RCT carried out longitudinal assessments, enabling the identification of the interventions’ long-term effects (20, 23, 25). Finally, the inclusion of a qualitative component was key to receiving participant and staff feedback, deepening the understanding of the program’s dynamics (12, 21).

Nonetheless, several weaknesses emerged across the studies. One of them is the absence of large-scale RCT, which undermines the rigor and generalizability of the findings. Many studies needed a control group or more data due to small sample sizes or drop-outs, limiting the ability to measure their effectiveness against traditional interventions (14, 22–23, 29). In addition to size, gender disparities appeared in study samples, composed mainly of women, which could influence the generalizability of the results (16, 22, 31).

Another weakness was the over-reliance on self-reported data, which introduces potential bias. Participants might not accurately report changes in their mental health due to cultural norms, stigma, or personal reservations (9, 32). Trust in subjective data weakens the reliability of the findings across all studies. Sadler, et al., identified the use of inadequate diagnostic tools as a limitation, underscoring the need to standardize evaluation methods to enhance the quality of the reviewed studies (15, 27).

Recruitment challenges were also a recurring issue. Many studies experienced difficulties in enrolling a sufficient number of participants due to cultural, social, or logistical barriers (26). High rates of people being left out and inconsistent participation, often because of health problems or scheduling conflicts, make the interventions less generalizable and strong (17, 25). Additionally, several studies pointed out the need for better follow-up after discharge, leaving uncertainty about long-term outcomes (18, 21, 24).

## Discussion

### Summary of evidence

The results highlight the potential of community-based interventions in reducing depressive symptoms in the elderly. These interventions can significantly improve their mental health and well-being. Our research found that interventions that include physical activity, social engagement, and mental health education have positively reduced depression. Including peer support groups or cognitive-behavioral therapy has demonstrated remarkable outcomes in alleviating depressive symptoms, improving social interaction and quality of life.

However, the variability in the effectiveness of the interventions discloses the need for more structured and standardized approaches. The community-based interventions have had notable success, some focusing solely on specific components, while others have integrated physical activities or cognitive behavioral therapy. The heterogeneity between the studies makes it challenging to compare them and conclude about which is the most effective intervention. Moreover, some factors like participant engagement, adherence, and the sociocultural environment also play a role in determining their success.

Sociocultural, financial, and geographical factors have influenced the implementation of community-based interventions. Local financial support and community engagement are some facilitators in helping get participation and improve outcomes, especially when tailored to contexts with specific needs (12, 19). However, there still exist cultural stigmas, geographical isolation, and logistical challenges in rural areas that keep posing barriers (13–14). Being able to address this requires a broad understanding of the sociocultural and geographical contexts, as well as innovative approaches to outreach and service delivery. By approaching this, health professionals can enhance the effectiveness and sustainability of community-based interventions, improving the well-being of vulnerable populations.

Across various interventions, a focus on integrating mental health education with social interaction is a common feature. Programs often combine group-based activities with educational sessions on mental health, like cognitive-behavioral techniques, empowering participants to manage their well-being proactively (17, 27). This combination of education and social engagement addresses emotional and practical aspects of mental health, providing participants with tools they can apply daily (12, 29).

However, several opportunities for improvement remain. Enhancing recruitment strategies by addressing barriers that hinder participation could improve the reach and impact of these programs (14, 26). Furthermore, providing personalized care for participants with severe mental health issues or cognitive impairments would make these interventions more inclusive and effective (9, 20). Lastly, improving accessibility by offering better transportation options or more flexible scheduling would ensure more consistent participation, enhancing the success of the interventions (12–13).

Community-based interventions have emerged as a promising strategy to address health outcomes effectively and affordably. A study found that they resulted in a significant decrease in the likelihood of rehospitalizations in patients with severe mental illness, which is critical for older adults who often have to face multiple health issues (33). Evidence suggests these interventions are inexpensive compared to traditional mental health treatments, which is a crucial consideration, especially in countries with limited resources. For instance, various studies have found a favorable incremental cost-effectiveness ratio (ICER) for community-based mental health services, ranging from $613 to $8,400 per quality-adjusted life year. This indicates that investing in community-based services can help improve health with benefits at a relatively low cost (33).

Community-based interventions, like the Atmiyata program in India, are cost-effective because they leverage local volunteers who provide mental health support without pay, reducing the need for expensive professional services. Additionally, these interventions utilize existing community resources and social capital, making them financially sustainable and scalable in resource-limited settings (34). This approach not only addresses mental health needs but also enhances community engagement and empowerment. Lay Health Workers (LHWs) is another strategy developed in Africa that reduces the need to hire highly specialized and expensive professionals. LHWs are often community members who receive specific training to carry out the interventions, reducing operating costs (35). Markedly, cultural adaptation enhances the acceptability and effectiveness of these interventions, reducing the need for repeated or additional ones and thus optimizing available resources (35). Hence, by addressing mental health problems early, these interventions can prevent more serious and costly long-term complications.

### Limitations

This research faced several limitations. Firstly, studies varied widely on the type and intensity of the interventions, the duration of follow-up, and the use of standardized measures. Outcomes were also heterogeneous among studies, mainly due to the absence of a clear consensus on how to screen for depression and measure improvements. The age limit that defines older adults also varied between studies. Many articles also suffered from methodologic limitations such as a small sample size or a lack of a control group, which limits generalizability. The identification of all relevant studies could have been reduced due to a deficiency in common taxonomy referring to community-based interventions and language limitations of the search, only including those spoken by the authors. Due to the wide array of studies retrieved in the search, the quality of individual sources of evidence was not evaluated. Finally, publication bias should be considered, as studies with unfavorable outcomes may not have been shared and disseminated.

### Recommendations for further research and practice

In this document, we identified several factors to be addressed in future research. We evidenced that most studies included Caucasian or Asian populations, disregarding Hispanic countries. Hence, there is an evident underrepresentation of Latin ethnicities, resulting in difficulties in extrapolating these interventions in Latin communities.

Community-based interventions are cost-effective strategies that improve well-being in areas where traditional healthcare services are deficient. However, many studies were conducted in urban settings, whereas few interventions were applied in rural and scattered areas, where these interventions are most effective, as they help bridge gaps in access to mental health care. Hence, more studies should detail the logistical strategies to achieve successful programs in dispersed and hard-access areas.

Additionally, there’s a lack of analysis concerning cost-effectiveness. Few studies included information regarding the funding and the cost of the interventions, limiting the application of these treatments in routine clinical settings. Future studies should include the practical and logistical considerations behind the implementation process for policymakers to fully evaluate their feasibility.

Although the short-term effectiveness of community-based interventions to address depression has been demonstrated in this review, their sustained effects through time remain controversial. Studies evaluating the maintenance of these effects in the long term should be developed, aiming to support the creation of public health policies backing their implementation.

### Conclusions

Community-based interventions for treating depression in older people have excellent qualities, such as focusing on social engagement, flexibility, doing thorough assessments, and being cost-effective. These programs effectively address the psychological and social challenges faced by older adults, fostering meaningful social connections and improving overall well-being. For future interventions to fully reach their potential, problems that currently exist need to be addressed, considering that rigorous randomized trials are limited, most of the data is based on self-reports, recruitment of participants is difficult, and tailoring the interventions to vulnerable groups faces many challenges. By refining these areas, these programs can become more effective in supporting the mental health and independence of elderly populations.

## Data Availability

All data produced in the present study are available upon reasonable request to the authors and are contained in the manuscript.

## Acknowledgement

Funding: This paper did not require external funding, with the exception of access to databases, which was covered by the subscriptions of the investigator’s institution.

## Author contributions

LMG and LRE conceptualized the research idea. LRE conducted the literature search, and LRE, MJ, IA, SS, VR, DU collected and analyzed the data. All authors contributed to the writing of the manuscript draft, reviewed and approved the final version of the manuscript.

## Appendices

**Appendix I:** Search conducted in four databases.

| Database | Search equation | Restrictions | Results |
| --- | --- | --- | --- |
| PubMed | ((("Aged"[Mesh] OR (elderly OR "older adults" OR "older people")) AND ((("Depression"[Mesh] OR "Depressive Disorder"[Mesh] OR "Depressive Disorder, Major"[Mesh]) OR (depres* OR "depressive disorder")))) AND ((("Community Mental Health Services"[Mesh]) OR ("community-based intervention" OR "community-based interventions" OR "community intervention" OR "community interventions")) | None | 821 |
| EMBASE | 'older people':ab,ti OR 'older adults':ab,ti OR 'aged'/exp AND 'depression'/exp OR 'major depression'/exp OR 'depres*':ab,ti OR 'depressive disorder':ab,ti AND 'community mental health service'/exp OR 'community-based intervention':ab,ti OR 'community-based interventions':ab,ti OR 'community intervention':ab,ti OR 'community interventions'.ab,ti | None | 173 |
| Ovid | exp Aged/ OR (elderly or "older people" or "older adults").ab,ti.<br>AND<br>exp Depression/ or exp Depressive Disorder/ or exp Depressive Disorder, Major/ OR (depres* or "depressive disorder").ab,ti.<br>AND<br>exp Community Mental Health Services/ OR ("community-based intervention" or "community-based interventions" or "community intervention" or "community interventions").ab,ti. | None | 94 |
| PsycNet | MeSH: Aged OR elderly OR "older people" OR "older adults"<br>MeSH: Depression OR MeSH: Depressive Disorder OR MeSH: Depressive Disorder, Major OR depres*<br>MeSH: Community Mental Health Services OR "community-based intervention" OR "community-based interventions" OR "community intervention" OR "community interventions" | None | 21 |

